# A Retrospective Analysis of Pediatric Mortality Trends Over Two Years at a National Referral Hospital in Mogadishu, Somalia

**DOI:** 10.1101/2025.07.05.25330946

**Authors:** Abdulkadir M Ahmed Keynan, Ahmed Mohamed dirie, Shafie Abdirahman Dirie, Mohamed Abdirahman Omar (Qalbi), Abdullahi Abdihakim Ahmed, Osman Abubakar Fiidow

**Affiliations:** Department Research, Training and Disease Surveillance at Banadir Hospital; Mogadishu Somalia; Dean Faculty of Health Science, Salaam University, Mogadishu, Somalia; Department of Community Health, Faculty of Medicine and Health Sciences, University Putra Malaysia, 43400 UPM, Serdang, Selangor, Malaysia; Somali national university and executive director at foolaad institute for health studies (FIHS); Department of nursing, faculty of health science, Salaam university, Mogadishu, Somalia; Department of Public Health, Faculty of Health Science, Salaam University, Mogadishu, Somalia; 2. Department of Research and Development, Somali Center for Research and Consultancy, Mogadishu, Somalia

**Keywords:** Retrospective, Demographic, Child, mortality

## Abstract

**Introduction:** Child mortality is a critical global health indicator, reflecting a nation’s development and healthcare quality. Despite significant progress in reducing child mortality globally, Somalia continues to experience high rates, particularly among infants and young children, due to preventable and treatable conditions. Malnutrition, birth complications, and infectious diseases are the leading causes of death. This study aims to analyze the demographic and mortality patterns of hospitalized children at Banadir Hospital in Mogadishu, Somalia, to identify key factors contributing to pediatric mortality and propose actionable solutions.

**Objective:** To assess and analyze the trends in pediatric mortality over the past two years at a national referral hospital in Mogadishu, Somalia.

**Design:** A retrospective cross-sectional study

**Setting:** Pediatric department at Banadir hospital in Mogadishu Somalia

**Participants:** 1,443 children admitted to the pediatric department

**Results:** The overall pediatric mortality rate during the study period was 5.3%. Male children accounted for 56.5% of deaths, while females represented 43.5%. Infants under one year were disproportionately affected, contributing to the majority of deaths. The primary causes of death were birth asphyxia (10.33%), severe acute malnutrition (29.04%), and preterm (13.65%). The majority of deaths occurred between two to five days and within the first 24 hours after admission, respectively.

Mortality was significantly associated with younger age groups, particularly neonates.

**Conclusions:** In order to reduce Somalia’s high rate of pediatric mortality, the results highlight the urgent need for better healthcare services and focused interventions. Enhancing maternal and neonatal healthcare, strengthening pediatric emergency care, and addressing malnutrition through community-based programs are critical. Increasing vaccination coverage and improving timely referral systems are also essential to prevent deaths from avoidable conditions. These strategies, if implemented effectively, can contribute to a substantial decline in preventable child deaths in Somalia.

## INTRODUCTION

Child mortality is a critical indicator that reflects a nation’s level of development(1,2). Child mortality has significantly decreased during the last 20 years. The under-5 death rate fell by 12% worldwide between 2015 and 2021, from 43 to 38 deaths per 1,000 live births. The global newborn mortality rate also decreased about 10%, from 20 to 18 deaths per 1,000 live births.(3) The Sustainable Development Goal (SDG) objective for under-five mortality had already been met by 133 nations as of 2021. However, 54 countries nearly 75% of which are in sub-Saharan Africa have not made enough progress, and more work is needed to reach the 2030 target.(4)

Globally, an estimated 26,000 children under the age of five die every day, with developing countries accounting for the majority of these deaths(2). In 2022, 4.9 million children under five died, including 2.3 million infants under one month and 2.6 million children aged one to 59 months (5). According to report from the World Health Organization (WHO), child mortality rates have decreased significantly, from 12.6 million in 1990 to 5.6 million in 2016 (6). Between 2000 and 2022, the global death rate for children under five dropped by 51% (5). Children aged 5-14 experienced the lowest mortality risk of any age group, with approximately one million deaths recorded in 2017 (7).In 2017, sub-Saharan Africa and Southern Asia were accounted for 50% and 30% of the global under-five mortality rates, respectively(4). According to previous studies on pediatric hospital mortality in Sub-Saharan Africa, high inpatient fatality rates, especially during early hospitalization, are caused by insufficient pediatric triage, assessment, and emergency care(2). This indicates the possibility of high mortality rates in Africa. In Sub-Saharan Africa, the infant mortality rate decreased from 182 to 58 deaths per 1000 live births between 1990 and 2017(9).

In Somalia, according to the Banadir Demographic Health Survey conducted in 2020, a significant number of maternal and neonatal deaths occur within the first 48 hours after delivery due to a lack of postnatal care.(10) The survey reported that only 8 percent of mothers received a postnatal checkup during this critical period (DHS). A study conducted in Mogadishu highlighted the high prevalence of premature births, which contribute to increased neonatal mortality.(11) Additionally, major challenges include the inadequacy and inaccessibility of medical services. the infant mortality rate dropped from 132 deaths per 1000 live births in 2000 to 80.4 and 73 deaths per 1000 live births in 2022, respectively(12). This suggests that the infant mortality rate in Somalia is among the highest in the world and the region. Malnutrition, including dietary deficiencies and inadequate breastfeeding, is a leading cause of morbidity and mortality among Somali children(13). 20–25% of all under-five mortality are caused by infections, including pneumonia and diarrhea, and delivery complications, which account for one-third of all neonatal deaths ^9^.

This study is the first of its kind in Somalia to our knowledge. Gathering some socio-demographic profile of hospitalized children, including age and sex, can help improve pediatric triage, optimize resource allocation, and enhance the quality of care.(14)

We aim to investigate child mortality among hospitalized children, considering socio-demographic factors, to identify the leading causes of death at the referral hospital in Mogadishu, Somalia.

## METHODS

### Study setting

This study was conducted at Banadir Hospital, a leading mother and child national referral and teaching hospital in Mogadishu the capital of Somalia. Operated by the Federal Ministry of Health (FMOH), the hospital is a cornerstone of pediatric healthcare in the region. It has a total capacity of over 700 beds, including 200 specifically allocated for pediatric patients, and is supported by a workforce of more than 501 healthcare professionals. The hospital provides a comprehensive range of pediatric services, including neonatal intensive care, pediatric emergency and critical care, a pediatric intensive care unit (ICU), and several inpatient wards. The pediatric emergency department handles all admissions for children who are referred to Banadir Hospital from other medical facilities, including as clinics, health centers, and other hospitals. Each year, the hospital admits approximately 12,000 pediatric patients, including those requiring specialized care in a neonatal unit for newborns under 29 days old. This extensive infrastructure and expertise position Banadir Hospital as a vital center for pediatric healthcare and education in the region.

### Study design and population

A two-year retrospective review was conducted to analyze the demographics and mortality of hospitalized children. The study utilized a convenience sampling technique, including all children admitted during the designated study period with complete and accurate documentation (i.e., no missing data). The target population comprised children recorded in the pediatric emergency register and the hospital inpatient ward register between January 1, 2022, and December 31, 2023. Children with incomplete documentation were excluded from the study.

In total, 1,513 children were admitted to the pediatric department during the study period. Of these, 70 patients were excluded due to incomplete records. Ultimately, 1,443 patients who met the study’s inclusion criteria were included in the analysis.

### Operational definitions

Neonate: A newborn child who is below 28 days of age(15)

AWD with severe: Acute watery diarrhea with Severe dehydration

Preterm birth: a newborn with gestational age is less than 37 weeks(16)

SAM with Complications: Severe acute malnutrition(17), Complications: like poor appetite, Hypoglycemia, hypothermia, dehydration and Anemia

### Data collection and data analysis

Data were collected using a structured form(1) designed to collect key variables age, gender, cause of death, date of admission, and date of death for all children documented in the pediatric hospital records, including the emergency and inpatient ward registers. The dataset was then analyzed using SPSS version 26 for statistical evaluation and Microsoft Excel for supplementary data management and visualization, ensuring a thorough and accurate analysis process. Descriptive statistics were utilized to present frequencies and percentages. The chi-square (χ^2^) test compared categorical variables related to causes of death and age groups. Data visualization included a line graph for mortality cases across quartiles and a bar graph for causes of death among children. The total number of pediatric admissions divided by the total number of hospital deaths was used to determine mortality rates.

### Patient and Public involvement

The public and patients could not be included in the planning, execution, and reporting of this research study.

## RESULT

During the two-year study period, which ran from January 2022 to December 2023, 27,076 pediatric patients were admitted to the hospital, and 1,443 of them died. This led to an overall child mortality rate of 5.3%. Table 1 displays the distribution of morbidity causes among 1,443 children across various age groups. A remarkable finding is that over half (58.7%) of cases with severe acute malnutrition (SAM) and complications occurred in children aged 1 to 5 years. Furthermore, nearly all cases of premature births (97.5%), birth asphyxia (96%), neonatal tetanus (100%), and neonatal sepsis (95.5%) affected children under one year of age. However, there was statistically significant association between the cause of mortality and the age group category (χ^2^=537.485, p<0.001).

**Table 1:** Morbidity causes across different age groups (n=1443)

| Cause of Morbidity | <1 year (n %) | 1-5 year (n %) | 6-10 year (n %) | >10 year (n %) |
| --- | --- | --- | --- | --- |
| SAM with Complication | 167(39.9) | 246(58.7) | 5(1.2%) | 1(0.2) |
| Premature | 192(97.0) | 0(0.0) | 4(2.0) | 1(0.5) |
| birth Asphyxia | 143(96.0) | 1(0.7) | 4(2.6) | 1(0.7) |
| Neonatal Tetanus | 87(100.0) | 0(0.0) | 0(0.0) | 0(0.0) |
| Measles with complication | 19(24.4) | 1(1.3) | 55(70.5) | 3(3.8) |
| Neonatal Sepsis | 63(95.5) | 0(0.0) | 3(4.5) | 0(0.0) |
| Severe pneumonia | 22(51.2) | 1(2.3) | 17(39.5) | 3(7.0) |
| AWD with severe dehydration | 11(27.5) | 1(2.5) | 27(65.5) | 1(2.5) |
| Meningitis | 16(42.1) | 1(2.6) | 16(42.1) | 5(13.2) |
| Shock | 8(32.0) | 1(4.0) | 15(60.0) | 1(4.0) |
| Aspiration | 12(38.7) | 8(25.8) | 6(19.5) | 5(16) |
| Anemia | 14(50.0) | 4(14.3) | 7(25.0) | 3(10.7) |
| Respiratory distress syndrome | 13(59.0) | 5(23.0) | 3(14.0) | 1(4.0) |
| Tuberculosis | 3(12.5) | 13(54.0) | 5(21.0) | 3(12.5) |
| Acute Kidney Injuries | 2(10.0) | 4(20.0) | 5(25) | 9(45) |
| Congenital Heart Disease | 10(55.6) | 5(27.8) | 2(11.1) | 1(5.5) |
| Gastroenteritis | 3(14.0) | 7(33.0) | 5(24.0) | 6(29.0) |
| Respiratory Failure | 5(26.0) | 4(21.0) | 7(37.0) | 3(16.0) |
| Neonatal Jaundice | 15(100.0) | 0(0.0) | 0(0.0) | 0(0.0) |
| Others* | 48(48.0) | 24(23.0) | 18(17.0) | 13(12.0) |

**\*Others:** Burn, Diphtheria, Low birth weight, Hydrocephalus, Cerebral palsy, Status epilepticus, Drug toxicity, Cellulitis, Leukemia and Whooping cough. $\chi^2 = 537.485$ , $P < 0.001$

The Distribution of Pediatric Deaths by Age and Sex is displayed in Table 2. Among deaths under one year, males accounted for 56% (506) compared to females at 43.5% (390). This trend persists across all age groups, indicating a consistently higher risk of mortality in males compared to females.

**Table 2:** Age & Sex Distribution of Pediatric Deaths (n = 1443)

| **Sex** | **>1 year (n %)** | **1-5 years (n %)** | **6-10 years (n %)** | **<10 years (n %)** | **Total Death (n %)** |
| --- | --- | --- | --- | --- | --- |
| **Female** | 390(43.5) | 211(43.9) | 18(40.0) | 9(42.9) | 628(43.5) |
| **Male** | 506(56.5) | 270(56.1) | 27(60.0) | 12(57.1) | 815(56.5) |
| **Total** | 896(100) | 481(100) | 45(100) | 21(100) | 1443(100) |

Figure 1 illustrates the prevalence of various causes of mortality among children. Severe Acute Malnutrition leads with the highest prevalence at 29.04%, followed by Premature at 13.65%. Shock has the lowest prevalence, standing at 1.733%. Based on the age groups, the mortality rates are highest among children under five years of age, particularly those under one year. Additionally, the majority of deaths in these age groups occur within the first 24 hours or between 2 to 5 days (Figure 2)

**Figure1:**
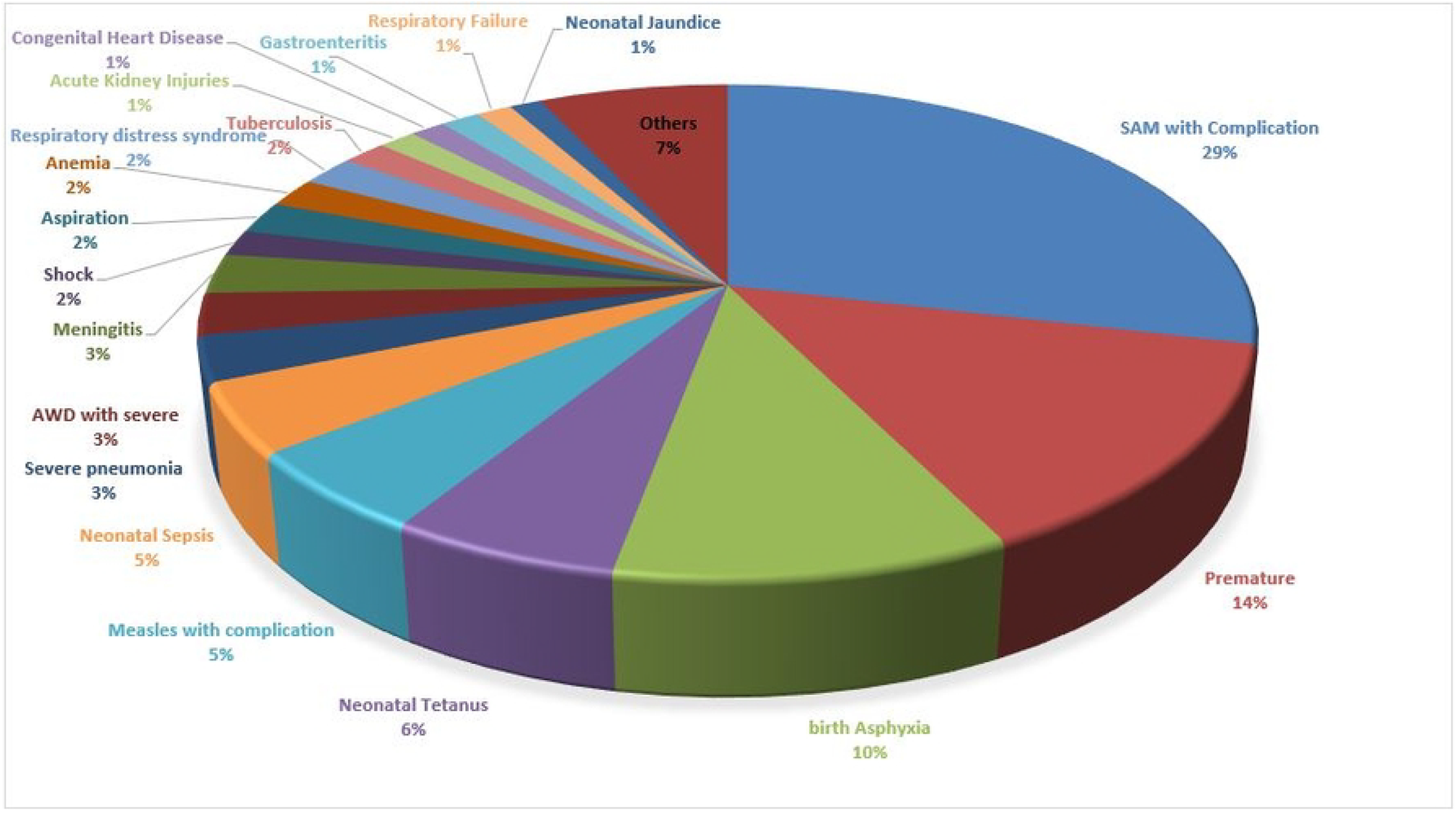
Causes of death among children

**Figure2:**
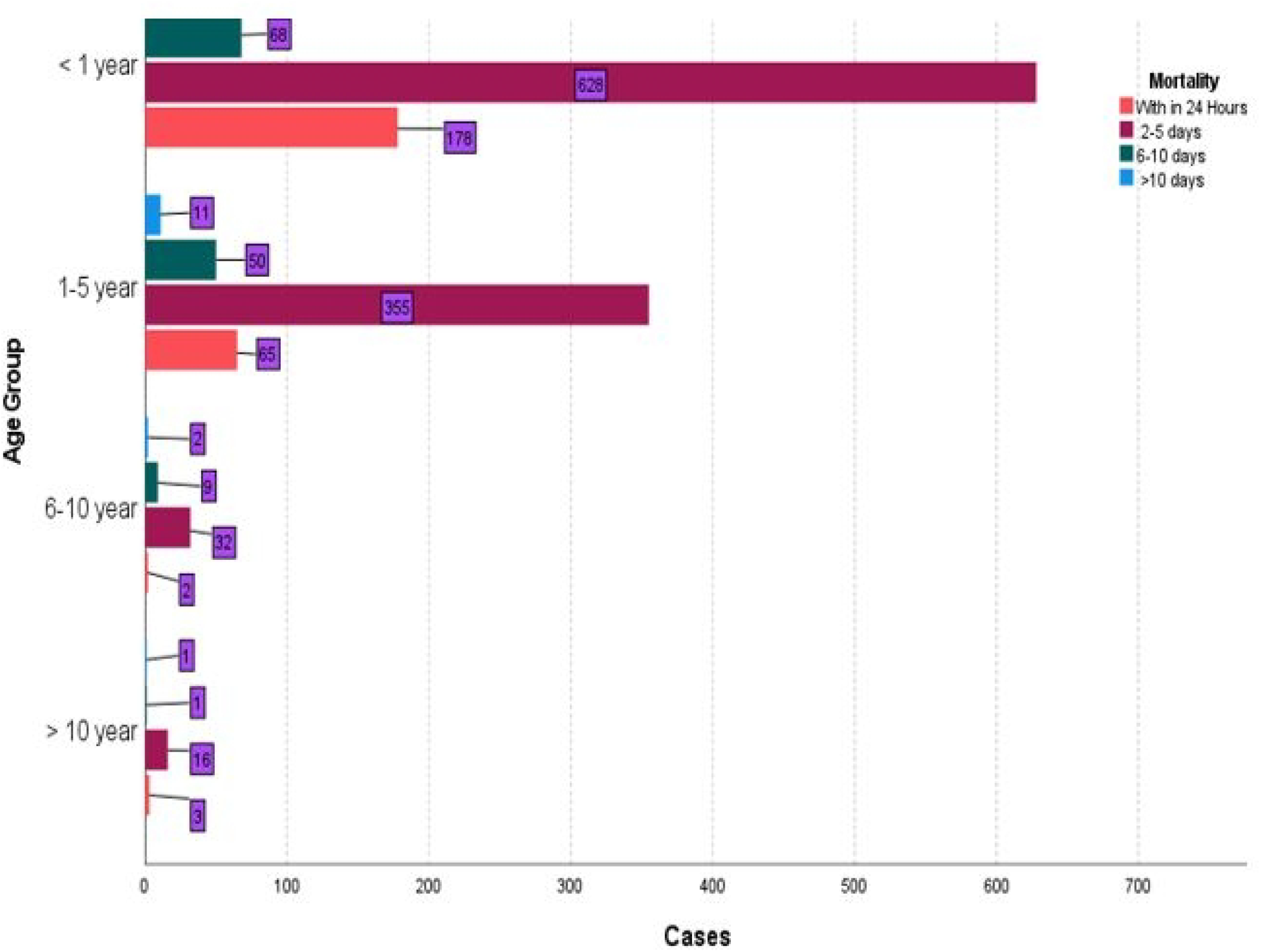
Age group and Death interval

Due to its significant contribution to mortality at all times, premature birth draws focus to the ongoing challenges with preterm babies. Between 2 and 5 days, the disease is responsible for 11.9% of cases, 19.4% during the first 24 hours, 30.6% after 10 days, and 11.7% from 6 to 10 days.

These numbers demonstrate how urgently premature children require specialized care facilities, early intervention, and all-encompassing neonatal support.

The primary cause of death at all intervals is SAM with Complications. Within the first 24 hours, it accounts for 19.0% of deaths, followed by 31.8% in the 2–5-day period, 28.1% in the 6–10-day period, and 22.2% beyond 10 days.

In the first 24 hours, birth asphyxia accounts for 16.5% of deaths, and in the next 2 to 5 days, it accounts for 9.6%. This emphasizes the significance of prompt newborn interventions and obstetric care in preventing asphyxia-related deaths, especially in high-risk deliveries. The fact that its percentage decreases after the first 24 hours suggests that birth-related actions can significantly lessen its effects. Table 3

**Table 3:** Interval days from the admitted day and the cause of death.

| Cause | 2-5 days (%) | 6-10 days (%) | >10 days (%) | Within 24 Hours (%) |
| --- | --- | --- | --- | --- |
| **Sere acute Malnutrition** | 328 (31.8%) | 36 (28.1%) | 8 (22.2%) | 47 (19.0%) |
| **Premature** | 123 (11.9%) | 15 (11.7%) | 11 (30.6%) | 48 (19.4%) |
| **Birth Asphyxia** | 99 (9.6%) | 8 (6.3%) | 1 (2.8%) | 41 (16.5%) |
| **Neonatal Tetanus** | 60 (5.8%) | 13 (10.2%) | 2 (5.6%) | 12 (4.8%) |
| **Measles with complication** | 61 (5.9%) | 9 (7.0%) | 1 (2.8%) | 7 (2.8%) |
| **Neonatal Sepsis** | 45 (4.4%) | 8 (6.3%) | 2 (5.6%) | 11 (4.4%) |
| **Severe Pneumonia** | 32 (3.1%) | 3 (2.3%) | 0 (0.0%) | 8 (3.2%) |
| **Acute watery diarrhoea with severe dehydration** | 27 (2.6%) | 1 (0.8%) | 0 (0.0%) | 12 (4.8%) |
| **Meningitis** | 27 (2.6%) | 5 (3.9%) | 1 (2.8%) | 5 (2.0%) |
| **Shock** | 18 (1.7%) | 3 (2.3%) | 0 (0.0%) | 4 (1.6%) |
| **Aspiration** | 19(61.3%) | 1(3.2%) | 0(0.0%) | 11(35.5%) |
| **Anemia** | 21(75.0%) | 4(14.0%) | 0(0%) | 3(11%) |
| **Respiratory distress syndrome** | 18(78.3%) | 1(4.3%) | 0(0.0%) | 4(17.4%) |
| **Tuberculosis** | 19(79.2%) | 2(8.3%) | 1(4.2%) | 2(8.3%) |
| **Acute Kidney Injuries** | 15(75.0%) | 0(0.0%) | 2(10.0%) | 3(15.0%) |
| **Congenital Heart Disease** | 12(63.2%) | 4(21.0%) | 1(5.3%) | 2(10.5%) |
| **Gastroenteritis** | 19(90.4%) | 1(4.8%) | 0(0.0%) | 1(4.8%) |
| **Respiratory Failure** | 11(57.9%) | 4(21.0%) | 0(0.0%) | 4(21.1%) |
| **Neonatal Jaundice** | 15(88.2%) | 0(0.0%) | 0(0.0%) | 2(11.8%) |
| **Others** | 87(74.4%) | 17(14.5%) | 8(6.8%) | 5(4.3%) |

### Mortality analyses based on Monthly

Figure 3 presents the monthly death counts for 2022 and 2023, highlighting the trends across the two years. In 2022, there was a significant spike in deaths in August, reaching a peak of 110, along with significant fluctuations throughout the year. In contrast, 2023 began with lower death counts, peaking at 88 in September.

**Figure3:**
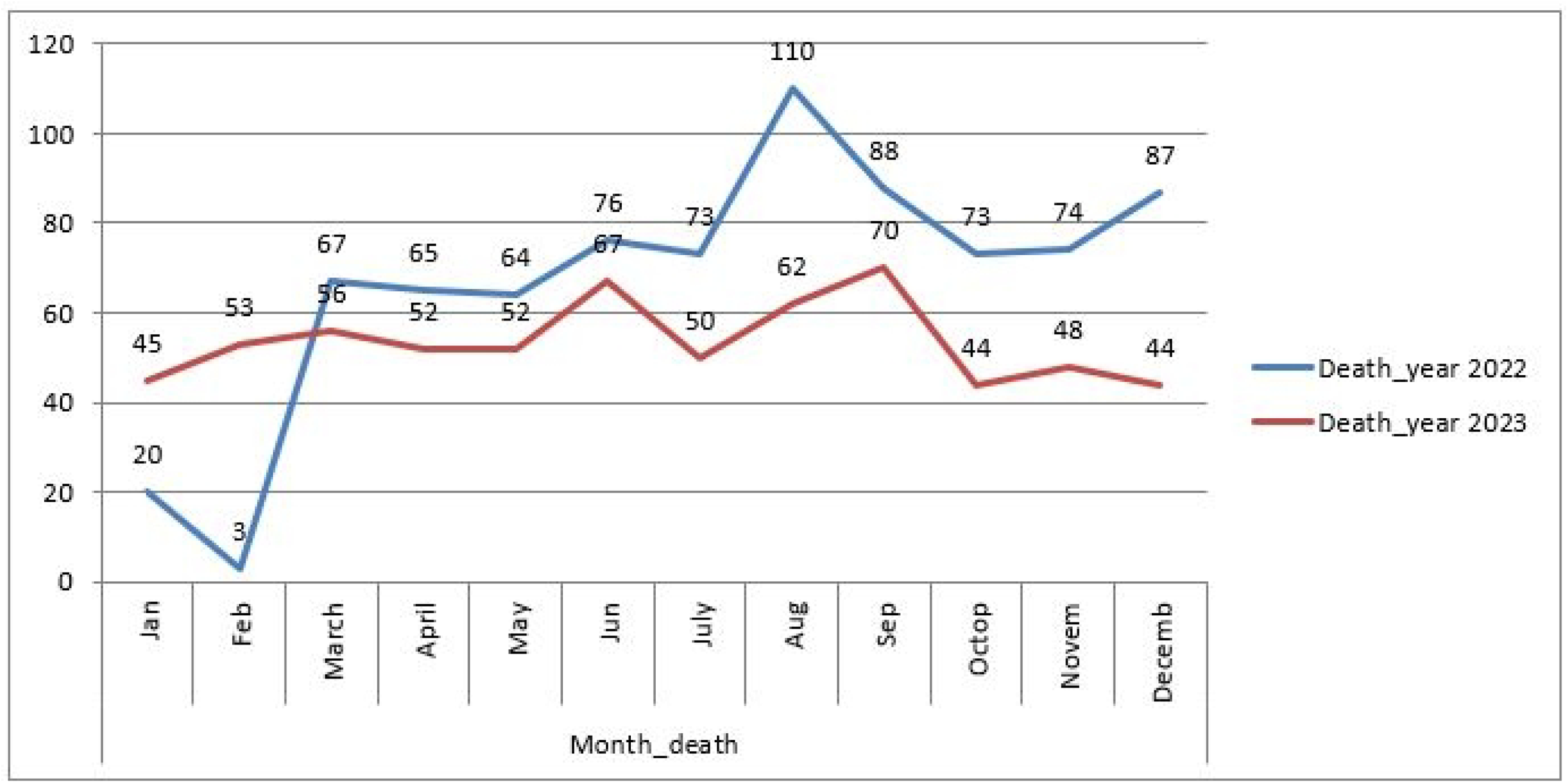
Mortality analysis based on Monthly

## DISCUSSION

Our Study highlights overall child mortality rate is 5.3%. This rate is higher than that reported in similar studies, including 4.57% at Latur Government Medical College and Hospital in India (19) and 0.042% at a referral hospital in Ethiopia (1). Despite this, the mortality rate that we found in the study is still lower than the reported national average for Somalia (20). The elevated mortality rate in our study may be due to a large proportion of critically ill admissions and delayed referrals of pregnant mothers and children to tertiary care hospitals.

Our study reveals that the majority of deaths occurred in the third quarter, followed by the fourth quarter, in both years when analyzed on a quarterly basis.(21) This pattern aligns with findings from other studies, which have shown that mortality rates are significantly influenced by seasonal variations, particularly during the third quarter. This trend can be attributed to several factors, including the **“hunger gap”** that coincides with the third quarter—a period when food stocks from the previous harvest are depleted, leading to widespread acute malnutrition. Additionally, Somalia frequently experiences drought conditions during this time, which severely impacts agricultural productivity and livestock health, both of which are vital for sustaining rural livelihoods. These combined factors contribute to the heightened mortality observed during this period.

Social determinants, including gender and age, significantly influence child and infant mortality rates(22).In our study, male children had higher mortality rates (56.5%) than females (43.5%) across all age categories. This is consistent with other research that indicated males are more likely than girls to die before the age of five (23). Generally, boys are more prone to premature birth and have a higher prevalence of congenital abnormalities and respiratory problems compared to girls (24).Male hospital admission rates were higher than those for female children (25). Also, it is worth note noting that our study participants were primarily male.

Infant (<1 year) mortality rates were highest compared to other age groups across both sexes, with males at 56.5% and females at 43.5%. This finding aligns with the previous study, which revealed the risk of death in the pediatric age group is highest during the neonatal period (19). One-third of all deaths in children under five occur in neonates, mostly as a result of infections and delivery complications (13). Our study found that the majority of deaths in these age groups occurred within the first 24 hours or between 2 to 5 days. This result is consistent with research done in Ethiopia, which reported that 50% of hospitalized children died within the first 24 hours of admission (14).Additionally, a systematic review by Sankar et al. (22) reported that a significant proportion of neonatal deaths occurred within the first three days of life, with two-thirds of these deaths happening on the first day alone.

SAM was the leading cause of morbidity in children aged 1-5 years (58.7%), followed by those under 1 year (39.9%). This aligns with findings from another study similar to those conducted in 95 different countries 17. Another study similar to our findings was conducted in the Kassena-Nankana District of Northern Chana, where malnutrition remains a major contributor to child morbidity due to food insecurity, inadequate healthcare access, and poor hygiene practices.

In the present study, birth asphyxia and prematurity emerged as the two most common causes of neonatal deaths, accounting for 10.0 % and 14.6% of all neonatal deaths, respectively. These findings are consistent with a study conducted in India (19) and also another study similar with our study conducted in Central Ethiopia (26)

In the current study, newborn tetanus was responsible for 6% of all deaths. This percentage is significantly lower than that seen in studies conducted in Nigeria, where a retrospective evaluation found a case fatality rate of more than 50%.(27)

Among the admitted children in the current study, measles contributed to 5% of all recorded deaths. This measles-related death rate is significantly more than what was found in a study done in Mogadishu, Somalia. Because Banadir Hospital is the biggest referral facility in Somalia, it receives the most severe and late-presenting cases from all over Mogadishu and beyond, which is why it records a higher number of measles-related deaths.(28)

Similarly, 5% of all deaths were due to newborn sepsis, which is less than the percentages found fin Ethiopian research.(29)

On the other hand, our results indicate that the most pronounced spike in mortality occurred in August and September. These findings are consistent with a study conducted in Kenya, which also reported a significant increase in mortality during the same period.(30)

While acute respiratory infections (ARI) are recognized globally as the leading cause of death in the pediatric age group, this study identified severe acute malnutrition, birth asphyxia, prematurity, measles, and neonatal tetanus as the primary contributors to child mortality.

Somalia faces distinct challenges contributing to higher mortality rates compared to global averages, highlighting the critical need for improved healthcare infrastructure, nutrition, and immunization efforts. Likewise, Banadir Hospital’s healthcare infrastructure is significantly under-resourced, with inadequate medical equipment, limited access to essential medications, a shortage of trained pediatric specialists and poor documentation. Despite its status as a referral hospital, these limitations restrict its capacity to effectively manage critically ill children, leading to increased mortality rates, particularly for severe cases.

Furthermore, the study’s limitations include the fact that only age and sex were examined as demographic factors because they were the only ones that were regularly noted in the hospital admission records. Additionally, septic shock and hypovolemic shock were also included in the study’s definition of “shock” according to the patients’ admission medical records.

### Conclusion

This study provides critical insights into the leading causes and demographic patterns of pediatric mortality at Banadir Hospital. However, due to the high rate of deaths within the first 24 hours of admission highlights the need of multifaceted approach which can greatly enhance neonatal outcomes including Increasing awareness and access to antenatal care (ANC), Strengthening hospital infrastructure, particularly by ensuring the availability of advanced medical technologies like mechanical ventilation and implementing comprehensive training programs for neonatal staff.

## ACKNOWLEDGEMENTS

We gratefully acknowledge the priceless assistance and guidance of the Banadir Hospital Research Center and the devoted staff of the Pediatric inpatient and out-patients without whom this work would not have been possible. Similarly, we extend our sincere appreciation to all authors for their significant contributions.

## Author Contribution

Conceptualization: AMAK, AMD. Data curation: AMAK, AMD. Formal analysis: AMAK, AMD, SHAD. Methodology: AMAK, Project Administration: AMAK, MAO. Resources: AMAK, MAO. Supervision: OAF, AAA. Validation: AMAK, AMD. Writing – original draft: AMAK, AMD, SHAD. Writing – review & editing: AMAK, AMD, OAF, MAO, AAA, SHAD. All authors have reviewed and given their approval for the final version of the manuscript.

## Guarantor

Dr. Abdulkadir M Ahmed Keynan is the guarantor of this work and accepts full responsibility for the integrity of the data and the accuracy of the data analysis.

## Data Availability Statement

The data of this study are available upon request from the corresponding author.

## Ethics statements

Consent of the patient for publishing

Not relevant

## Ethics Approval

The study was approved by the Ethics and Research Committee of Somali National Institute of Health under approval reference number (Ref: NIH/IRB/18/FEB/2025), and permission of hospital administration. All collected data were handled with strict confidentiality, ensuring no individual-level information was disclosed. The results will be presented in aggregate form to protect participant privacy and adhere to ethical guidelines.

